# The effectiveness and safety of high-intensity interval training, yoga and intermittent hypoxia-hyperoxia exposure on cognitive performance and health in individuals with mild cognitive impairment (KAYH): a study protocol for a randomized, sham-controlled trial

**DOI:** 10.64898/2026.08.05.26359785

**Authors:** Silas Wagner, Daniel Haigis, Mirela Bilc, Eva Beiner, Andreas M. Nieß, Andreas J. Fallgatter, Gerhard W. Eschweiler, Inga Krauß, Holger Cramer

**Author notes:** Correspondence: Silas Wagner, University Hospital Tübingen, Department of Sports Medicine Address: Hoppe-Seyler-Str. 6, 72076 Tübingen, Germany Mail. These authors contributed equally to this work and share last authorship (Inga Krauß, Holger Cramer).

## Abstract

**Introduction:** Individuals diagnosed with mild cognitive impairment (MCI) have an increased risk of developing dementia. Since there are currently no curative pharmacological therapies for MCI, nonpharmacological and exercise-related medical therapies represent a promising approach. This study aims to evaluate the effects of nonpharmacological interventions on cognitive performance in individuals with MCI.

**Methods and analysis:** The study will be a prospective, randomized, sham-controlled, single-centre superiority trial with a parallel-group design. A total of 100 patients aged ≥ 60 years with MCI will be randomly assigned to one of the three intervention groups or the control group. The three intervention arms comprise high-intensity interval training (HIIT), yoga, and intermittent hypoxia-hyperoxia exposure (IHHE), whereas participants in the control group will receive a sham application of IHHE (IHHE-S). The primary outcome is the cognitive function after 3-month intervention period assessed by Montreal Cognitive Assessment. The secondary outcomes and the evaluation of modifiable risk factors for dementia include quality of life, laboratory data, physical activity, anthropometric data, and cardiorespiratory fitness. All harms and (serious) adverse events will be assessed systematically at each study visit.

**Ethics and dissemination:** This study has been approved by the Ethics committee of the Medical Faculty of the University of Tübingen (494/2025BO1). Research findings will be published in peer-reviewed journals and presented to stakeholders and at scientific conferences.

**Trial registration:** The study was registered on the German Clinical Trial Register DRKS on 04.12.2025 (DRKS00038438).

**Strengths and limitations of this study:**

- The randomized, sham-controlled study design represents a key methodological strength, allowing reliable comparisons between the intervention groups and the control group while accounting for potential placebo effects.
- The 12-month follow-up from baseline provides the opportunity to evaluate the long-term effects of the interventions.
- Blinding of participants and therapists cannot be fully implemented across all study groups because of the nature of the interventions.

## Introduction

Due to demographic change and the resulting aging of the population, the provision of care for individuals with age-associated diseases is becoming increasingly important. Among those aged 60 years and older, the most common conditions are neurocognitive disorders, such as dementia. Dementia is a decline in cognitive function that affects memory and thinking, interfering with a person’s ability to function independently in their daily life [1, 2]. Ensuring optimal care and social participation for people with dementia places considerable demands on both the healthcare system and society [3]. An international working group projects a substantial rise in dementia prevalence, predicting a threefold increase in the number of individuals affected worldwide over the next three decades. Specifically, the estimated number of global cases is expected to increase from 57.4 million in 2019 to 152.8 million in 2050 [4]. In Germany, the number of people with dementia was estimated to be approximately 1.8 million in 2021 [3]. Projections indicate a potential increase to 2 million people aged 65 or older with dementia by 2033 [3] and approximately 2.8 million by 2050 [4]. This rapid increase underscores the urgent need for preventive measures on the basis of robust scientific evidence.

Mild cognitive impairment (MCI) often is a prodromal stage of dementia. MCI is characterized as the stage between normal age-related cognitive changes and dementia, and is typified by detectable cognitive decline without any impact on activities of daily living [5, 6]. Diagnosing MCI offers the opportunity to prevent subsequent severe cognitive or dementia-related conditions and allows for effective secondary prevention strategies [7]. According to international data, the global prevalence of MCI among people aged 50 years and over is approximately 19.7% [8]. A recent meta-analysis of 89 studies reveals that, for patients with MCI, the risk of converting to dementia is 41.5% in clinical settings and 27.0% in population-based settings during a mean follow-up period of 5.2 years [9]. Moreover, the reversion rates to normal cognition are 8.7% and 28.2% in these settings, respectively. This highlights the urgent need for the treatment of MCI in individuals aged 60 years and older, ultimately aiming at dementia prevention. As no curative pharmacological interventions currently exist for MCI or dementia, nonpharmacological therapeutic approaches represent promising options. Various nonpharmacological interventions have been investigated in this context [10, 11]. Nevertheless, the exact effects in terms of efficacy, safety, and feasibility remain only partially understood.

High-intensity interval training (HIIT) is a specific form of endurance training, that alternates short periods of high-intensity exertion with recovery phases. Initial evidence from a review by Marriott et al. (2021) suggests that HIIT offers greater health benefits than continuous moderate-intensity endurance training and is generally well tolerated by older adults [12]. A recent meta-analysis suggests that HIIT can be an effective intervention to improve cognitive function in older adults and patients with cognitive impairment [13]. Furthermore, a review examining high-intensity training in individuals with MCI or dementia, including various training modalities (endurance, functional, and resistance training), reports that high-intensity training generally improves cognitive function, or at least delays global cognitive decline [14]. Analysis of the three studies included in the review, which examined the effects of high-intensity endurance training, reveals improvements in global cognitive function in individuals with MCI [14]. However, owing to the limited number of studies, small sample sizes, and substantial heterogeneity among trials, the current evidence is insufficient, and further research into the efficacy, feasibility, and safety of HIIT in MCI or dementia is needed.

For individuals with MCI, mind-body therapies such as yoga may also represent promising options in terms of efficacy, safety, and feasibility [15, 16]. Earlier systematic reviews of yoga-based interventions in healthy older adults report improvements in cognition [17, 18], balance, flexibility, muscle strength, and depressive symptoms [19], as well as increased health-related quality of life and psychological well-being [20]. In terms of safety, previous studies report that yoga is safe for older adults, including those with MCI. In mixed samples of older adults without cognitive impairment as well as those with MCI and dementia, Bhattacharyya et al. (2021) report moderate positive effects of yoga on cognitive function [21], with effect sizes comparable to those of cholinesterase inhibitors [22]. While these findings are promising, they also underscore the need for high-quality randomized controlled trials to confirm the efficacy of yoga for improving cognitive health and to elucidate the underlying mechanisms.

Another potential nonpharmacological and noninvasive approach for the treatment and prevention of neurodegenerative and neurovascular diseases is intermittent hypoxia exposure (IHE) [23]. To enhance the effects of IHE, replacing the normoxic phases with hyperoxia during intermittent hypoxia-hyperoxia exposure (IHHE) may improve the impact of hypoxia [23, 24]. IHHE is generally assumed to share similar mechanisms of action with IHE [25]. A recent meta-analysis of cognitive outcomes suggests that IHHE may be an effective intervention for improving physical and cognitive performance and reducing cardiometabolic risk factors in older adults with cardiovascular and metabolic diseases or cognitive impairment [26]. Several systematic reviews show that IHHE is safe and well tolerated in both younger and older individuals [27, 28]. However, further high-quality randomized controlled trials are needed to confirm these potential benefits and strengthen the evidence base [26].

## Objectives

The primary aim of this study is to evaluate the effectiveness, safety, and feasibility of three different non-pharmacological interventions from the domains of exercise therapy (HIIT), mind-body practice (yoga), and physical therapy (IHHE) on cognitive and physical performance and health in individuals with MCI compared to a sham application of IHHE (IHHE-S). The primary outcome measure will be cognitive function after 3 months of intervention.

The primary hypothesis of this trial is:

1. Each of the three active intervention arms is superior to the control group in global cognitive function after 3 months of treatment.

Secondary hypotheses are:

2. Each of the active intervention arms is superior to the control group for secondary outcomes after 3 months of treatment.
3. Superiority of the three active treatment arms for global cognitive function and patient-reported secondary outcome measures remains for another 9 months follow-up after the end of intervention.

## Methods and analysis

### Study design

This is a prospective, randomized, sham-controlled, single-centre superiority trial with a parallel-group design. The trial is designed to demonstrate the superiority of three active interventions over sham with respect to the primary outcome, global cognitive function. The unit of randomization is the individual participant and the participants will be allocated in a 1:1:1:1 ratio to one of three active intervention groups or to the sham group. The study will be conducted at the University Hospital of Tübingen, a tertiary care centre in Baden-Württemberg, Germany, and will be organized with continuous study inclusion over a period of approximately one year. Recruitment will primarily take place in outpatient memory clinics and specialty outpatient units at the University Hospital of Tübingen. This protocol is developed in accordance with the Standard Protocol Items: Recommendations for Interventional Trials (SPIRIT) guidelines 2025 [29] and the SPIRIT 2025 explanation and elaboration [30]. The SPIRIT checklist is presented in Supplementary 1.

### Study population

The eligibility criteria were defined to ensure the recruitment of a representative cohort of patients with MCI while minimizing potential confounding factors and ensuring participant safety. All participants must meet all inclusion criteria and none of the exclusion criteria (see Table 1).

**Table 1:** Inclusion and exclusion criteria of the KAYH study.

| Inclusion criteria | Exclusion criteria |
| --- | --- |
| - MCI (classification according to DemTect < 13 [cut-off for cognitive impairment] and DemTect > 8 [cut-off for no dementia]; existing MCI diagnosis; DemTect score of 13–15 and SLUMS score of 21–26)<br>- Physical condition that permits the use of a cycle ergometer and participation in the interventions<br>- Sufficient time availability (e.g. assessments, intervention)<br>- Adequate language, visual, and hearing ability for completion of self-reported and interviewer-administered assessment instruments<br>- Ability to travel to the study center<br>- Age ≥ 60 years<br>- Provision of informed consent and willingness to participate in the study | - Severe psychiatric and neurological disorders (e.g., Parkinson's disease, Huntington's disease, schizophrenia, bipolar disorder, cerebrovascular disease with acute stroke or cerebral hemorrhage within the past 6 months)<br>- Congenital or acquired cardiac and major vascular malformations<br>- Renal failure<br>- Diagnosed dementia<br>- Comorbidities that contraindicate physical exercise, HIIT, Yoga or IHHE (e.g., untreated hypertension, coronary artery disease with unstable angina pectoris [Canadian Cardiovascular Society, CCS grade 3–4], heart failure [New York Heart Association, NYHA class II–IV], myocardial infarction within the past 3 months, pulmonary fibrosis, chronic obstructive pulmonary disease [GOLD stage II, FEV<sub>1</sub> < 80% predicted at rest], bronchial asthma with significant dyspnea, oncological diseases in the acute phase/acute treatment, acute inflammatory or somatic diseases)<br>- Abnormal findings during baseline (t0) ergometry that represent a contraindication for HIIT, Yoga or IHHE (e.g., musculoskeletal complaints or clinically relevant ECG or blood pressure abnormalities)<br>- Requirement for supplemental oxygen (continuous or intermittent, device-assisted)<br>- Resting oxygen saturation < 90%<br>- Individual intolerance or hypersensitivity to oxygen<br>- Current smoking<br>- Alcohol dependence within the past 2 years<br>- Multiday stay at altitudes > 1,500 m above sea level within the past 4 weeks<br>- Current yoga practice<br>- ≥ 150 minutes of moderate or ≥ 75 minutes of vigorous physical activity per week (excluding daily activity for transportation purposes)<br>- Concurrent participation in another clinical intervention study |

Notes: DemTect: Dementia detection test; SLUMS: Saint Louis University Mental Status; MCI: Mild cognitive impairment; HIIT: High Intensity Interval Training; IHHE(-S): Intermittent hypoxia-hyperoxia exposure (-sham).

### Recruitment

Physicians at the University Hospital Tübingen will approach potentially eligible patients in outpatient clinics of the University Hospital or following inpatient stays (e.g., gerontopsychiatric outpatient clinic, memory clinic, inpatient treatment in the Department of Psychiatry and Psychotherapy, and inpatient treatment in cooperating facilities such as the stroke unit) and will provide them with information about the study (e.g., informational flyer). Further recruitment strategies refer to regional information events on dementia (e.g., Alzheimer Society Baden-Württemberg), articles and press releases in the local newspapers as well as via social media accounts and the homepage of the University Hospital Tübingen. If interested, participants contact the study team at the University Hospital by email or phone. The initial screening phone call then serves to inform potential participants on study contents, aims and the timeline of the study. Subsequently, potential eligible and interested patients will be invited to a screening visit, during which their eligibility will be assessed according to the predefined inclusion and exclusion criteria. The research team, comprising physicians and psychologists, will provide potential participants with detailed information about the study and obtain written informed consent. Following confirmation of eligibility and consent, participants will be provisionally enrolled in the study. At this stage, all inclusion and exclusion criteria, except for contraindications identified during physical examination and ergometry, will be assessed. Final study inclusion by a study physician will occur after ruling out contraindications during the baseline medical examination (t0) conducted in the Department of Sports Medicine, University Hospital Tübingen, contingent upon normal cardiopulmonary exercise testing (spiroergometry) and medical clearance for exercise. If, after provisional inclusion, a participant is excluded during this process, the case will be classified as a screening fail-out, without final inclusion or randomization.

We plan to recruit participants and include them in the study continuously over a period of approximately one year. The recruitment rate will fluctuate depending on factors such as irregular special events and recruitment information sessions. On average, we expect a recruitment rate of three participants per week.

### Intervention and comparator

All interventions will be conducted over a period of 12 weeks, involving two sessions per week, with each session lasting approximately 45 minutes. The detailed dosage criteria including exercise frequency, intensity, time and type (FITT), for all interventions are presented in Table 2. The three active interventions are described according to the Template for Intervention Description and Replication (TIDieR) checklist and guide [31] and the sham-intervention is described according to the TIDieR-Placebo guide and checklist for reporting placebo and sham controls [32]. The respective checklists are presented in Supplementary 2.

**Table 2:** Detailed FITT criteria for all interventions.

|  | **HIIT** | **Yoga** | **IHHE** | **IHHE-S** |
| --- | --- | --- | --- | --- |
| **Frequency** | 2 sessions/week | 2 sessions/week | 2 sessions/week | 2 sessions/week |
| **Intensity** | Week 1-4: 50-60% VO<sub>2</sub>peak; RPE: 12-13<br>From week 5: 70-79% VO<sub>2</sub>peak; RPE 15-16, possible increase up to >80% VO<sub>2</sub>peak, RPE 17-18) | Group-oriented depending on individual performance | Individualized after initial hypoxia testing 10-14%/30-40% FiO<sub>2</sub> | Normoxia at ambient air ~21% FiO<sub>2</sub> (sham-IHHE) |
| **Time** | 10' warm-up; 25' HIIT (4 sets (4' interval, 3' active break at 40-60% HR<sub>max</sub>); 10' cool-down; total time ~45' | 45' | 4-8 cycles with hypoxia for 1-3,5' and hyperoxia for 1-6'; total time ~45' | Normoxia for ~45' |
| **Type** | Interval training | Hatha yoga (postures, breathing, and meditation techniques) | Intermittent | Constant |
| **Modality** | Cycle ergometer | Group course led by certified yoga instructors | ReOxy; Ai Mediq, (Luxembourg) | ReOxy; Ai Mediq, (Luxembourg) |
| **Setting** | Department of Sport medicine | University Hospital of Psychiatry and Psychotherapy | Department of Sport medicine | Department of Sport medicine |
| **Supervision** | 1:4 | 1:8 | 1:2 | 1:2 |

Notes: HIIT: High Intensity Interval Training; IHHE(-S): Intermittent hypoxia-hyperoxia exposure (-sham). minutes: ' ; HR<sub>max</sub>: maximum heart rate; HRR: Heart Rate Reserve; VO<sub>2</sub>peak: maximum oxygen uptake; RPE: Rating of perceived exertion (6-20); FiO<sub>2</sub>: fraction of inspired oxygen.

The individuals responsible for conducting the interventions are required to meet specific prerequisites. For instance, yoga classes are exclusively delivered by certified yoga instructors, while the other interventions are facilitated by trained sport and exercise scientists. All persons carrying out the interventions will be trained accordingly. It is foreseen to pilot the HIIT, IHHE and IHHE-S interventions in advance as part of a pilot project.

### High intensity interval training (HIIT)

The training protocol applied in this study is based on a previous investigation into the efficacy of HIIT in individuals with MCI and hypertension [33], as well as on position papers and reviews on HIIT in older adults and individuals with cardiac risk factors [14, 34]. Training will be performed on a cycle ergometer and monitored via power output in watts and continuous heart rate recording. In addition, perceived exertion will be recorded after each interval via the rating of perceived exertion (RPE) scale. The training protocol incorporates a 4-week familiarization phase of moderate continuous endurance training to establish a basic fitness level and to physically and mentally prepare participants for interval training [35]. During the 4-week preparatory phase, following a 10- minute warm-up, the intensity will be increased to achieve a target moderate cardiovascular load and maintained for 30 minutes, followed by a 5-minute cool-down. HIIT commences from the fifth week onwards. After a 10-minute low-intensity warm-up, the main training phase will consist of four 4-minute high-intensity intervals, interspersed with 3-minute active recovery periods. The session will conclude with a 10-minute cool-down, totaling approximately 45 minutes. Training sessions will be supervised in the Department of Sports Medicine at a maximum instructor-to-participant ratio of 1:4 by trained personnel with a degree in sport and exercise science.

### Yoga

The yoga intervention will be delivered as a group course with a maximum of 8 participants per group. Over a 12-week period, three 45-minute classes per week will be offered, with participants instructed to attend only two sessions per week. The course will be led by qualified yoga instructors and will consist of a combination of physical postures (asanas), relaxation and meditation (dhyana), and breath regulation techniques (pranayama), adapted to the target population. Within the group courses, the content will be tailored by the yoga instructors as much as possible to the individual circumstances of the participants and may be adjusted during the single intervention sessions. For this purpose, a manual for a 12-week yoga course was developed, which allows entry at any time due to the continuous study enrollment. The content of the yoga therapy program was developed on the basis of systematic literature searches and the expertise of an interdisciplinary team with extensive experience in sports therapy, integrative medicine, and clinical care.

### Intermittent hypoxia-hyperoxia exposure (IHHE)

Before the first regular session and then every four weeks thereafter, participants will perform a continuous hypoxia test via the IHHE device (ReOxy; Ai Mediq, Luxembourg, Class IIb medical device), during which they will breathe a gas mixture containing 12% fraction of inspired oxygen (FiO) through a face mask for a maximum of 10 minutes. During both the test and all subsequent IHHE sessions, participants will be seated in an armchair, and heart rate as well as peripheral oxygen saturation (SpO) will be monitored continuously via pulse oximetry, with SpO not permitted to fall below 75% (target range 85–90%). Additionally, the subjects’ blood pressure will be recorded before and after each application. On the basis of the individual response during the hypoxia test (time to reach target SpO and time to recovery of baseline SpO), the device algorithm will determine personalized IHHE treatment parameters [23, 26]. Each IHHE session will comprise 4–8 cycles of hypoxia and hyperoxia lasting 1–3.5 and 1–6 minutes, respectively. The oxygen concentrations in the hypoxic and hyperoxic phases (10–15% and 30–40% FiO, respectively) are automatically regulated by the IHHE device. The total session time, including preparation and recovery, will be approximately 45 minutes.

### Control group (IHHE-S)

A sham comparator was chosen to assess the intervention’s efficacy and safety while controlling for potential placebo effects. This ensures that any observed effects can be attributed to the three active interventions rather than to participant expectations or other nonspecific factors. The use of a sham control is consistent with ethical guidelines, as there is no established therapy that would preclude its use in this study population. The sham application of IHHE (IHHE-S) was chosen as the control group because, of the three active interventions, IHHE is the only one for which blinded sham control is possible. Previous studies on IHHE in individuals with MCI have used the IHHE-S as a control group and thus offer comparability [36, 37].

Participants in the control group will receive IHHE-S sessions twice weekly for 12 weeks. Similar to the IHHE intervention group, a 10-minute sham hypoxia test under normoxic conditions will be performed prior to the first regular session and then every four weeks thereafter. Subsequent IHHE- S sessions, including preparation and recovery, will be conducted under normoxic conditions (with ambient air at 21% FiO) for approximately 45 minutes [23, 26, 37]. Participants will be seated in an armchair during the session, and heart rate and SpO will be continuously monitored and blood pressure before and after each session will be recorded. Both the IHHE and IHHE-S sessions will be supervised in the Department of Sports Medicine at a maximum instructor-to-participant ratio of 1:2 by trained sport and exercise scientists.

The intervention discontinuing criteria include participant request to withdraw, medical reasons or the occurrence of (serious) adverse events ((S)AEs) attributable to the intervention. No modifications, such as adjusting session frequency or duration, will be made to the intervention protocol. However, single intervention modalities can be adapted to participants and their individual responses (e.g., intensity for HIIT with RPE), as described above. These changes will be documented in the study records.

During each therapy session, participants’ current state is assessed immediately before and after the session. In addition to the Feeling Scale [38], participants are asked one question each about their current energy level, general mood, physical discomfort or pain, and fatigue/recovery, using a 5- point Likert scale. The perceived effort or strain of the intervention is also assessed using the RPE scale. As part of the yoga intervention, two additional questions about specific side effects are asked after each session. During the first and last therapy sessions, participants’ expectations and evaluations of the therapy are also assessed [39].

Adherence to the intervention will be continuously tracked by recording attendance at all scheduled training sessions and by monitoring compliance with prescribed exercise intensities (e.g., via heart rate monitoring, rating of perceived exertion or pulse oximetry). Participation in yoga classes will be recorded by the yoga instructor. During the individual sessions, adherence to the prescribed modalities is ensured through the supervision and monitoring of the trained instructors. Any modifications or deviations from the established HIIT and IHHE protocols, such as adjustments to target heart rate zones, or deviations from the pre-defined yoga protocol will be systematically documented. To improve adherence, participants will receive notes detailing their intervention session appointments to help them remember to attend.

Participants will be advised to continue standard medical care as prescribed by their primary healthcare providers, and deviations will be recorded to assess the potential impact on the study results. Participants are not permitted to engage in any of the other intervention forms during the course of their involvement in the intervention, except for the one assigned to them. Conversely, other forms of therapy or concomitant care, such as cognitive training, may be administered without additional restrictions. All concomitant treatments and therapies that participants receive during the study period will be documented.

### Sample size

The sample size calculation was performed using the Montreal Cognitive Assessment (MoCA) [40]. According to previous studies, a group difference of at least two MoCA points can be considered minimally clinically significant [41]. Assuming a standard deviation of 2.1 points corresponding to the MCI population [42] and an assumed group difference of 2 points in the MoCA, the effect size is d=0.952 or f = 0.476. Based on this effect size, an analysis of covariance with the intervention group as the between-subjects factor (four groups), baseline as the covariate and a two-sided significance level of 5% and power of 95%, the required sample size is 80 patients. Considering a dropout rate of 20%, 100 subjects will be included in the study.

### Randomisation

The participants will be randomly assigned to one of the three intervention groups or the control group at a ratio of 1:1:1:1. A computer-generated randomization list will be created prior to study initiation by using R software and Excel, and implemented on the REDCap (Research Electronic Data Capture) platform. The randomization sequence will be prepared by an independent researcher not otherwise involved in participant enrollment or treatment. Participants will be randomized using a restricted randomization procedure with block randomization to ensure balanced group sizes throughout the enrollment period. Blocks of randomly varying sizes (4, 8 or 12 participants) will be used to make the allocation sequence less predictable. No stratification will be performed. Those responsible for enrolling participants will not have access to the randomization list, and all relevant fields in REDCap will remain hidden until randomization has been performed. Only the researcher responsible for generating the randomization list will have access to it. Study personnel involved in participant enrollment will not have access to the randomization list or the corresponding allocation fields within REDCap. Allocation will be performed automatically by the REDCap system immediately after enrollment is complete, ensuring concealment of allocation.

### Blinding

Blinding of the intervention facilitators is not feasible. After randomization, group assignment will only be visible to authorized personnel of the study team in REDCap. Furthermore, the baseline assessment is performed before allocation and is therefore kept blinded. For the primary endpoint, blinding of the outcome assessors is intended and data analysis of the primary outcome will be conducted in a blinded manner. Assessment of the primary outcome at follow-up will be performed by physicians or researchers who are not involved in the delivery of the intervention and who will therefore be blinded. Accordingly, the data analysis of the primary outcome will also be performed by a researcher who is not involved in the recruitment, assessment or delivery of the intervention.

The IHHE and IHHE-S are blinded to the participants, as the two intervention arms always take place at different time points but in the same room and use the same device, equipment and settings. Furthermore, participants will not be able to see the IHHE device display. A blinding assessment will be performed on the IHHE and IHHE-S groups at follow-up t1 to evaluate the success of the blinding process [43]. The participants in the HIIT and yoga interventions cannot be blinded. No reasons are anticipated that would justify unblinding for the IHHE and IHHE-S groups. However, if this were to occur, the principal investigators will have access to group allocations, and any unblinding would be reported.

### Outcomes Baseline variables

During the screening process, the Dementia-Detection test (DemTect) will be administered as a cognitive screening instrument to identify participants meeting the inclusion criterion of MCI [44]. In addition, the Saint Louis University Mental Status (SLUMS) examination is used in the screening process [45]. At baseline, comorbidities, concomitant care and current medication will be documented, and sociodemographic variables, including age, sex, ethnicity, education level, employment status, and living situation, will be collected. Any new comorbidities, concomitant treatments or medication changes will be recorded at subsequent measurement points.

We will assess modifiable risk factors for dementia. The potentially modifiable risk factors for dementia are based on a recent review by Livingston et al. (2024) [11]. The risk factors listed are low education, hearing loss, high LDL cholesterol, depression, traumatic brain injury, physical inactivity, diabetes, smoking, hypertension, obesity, excessive alcohol, social isolation, air pollution, and visual loss [11]. All of these risk factors, except for air pollution, will be recorded within our study. As previously described, educational level, hearing and visual loss, traumatic brain injuries, and smoking and alcohol consumption habits are assessed as baseline variables. All other risk factors are recorded as part of patient-reported outcome measures (PROMs) and the collection of secondary outcomes, and are described below.

### Primary outcome measures

The primary outcome will be global cognitive function, assessed by the (MoCA) [40]. The primary analysis will focus on the mean change in the MoCA score from baseline to 3 months post-randomization (t1).

### Secondary outcome measures

Visual attention and executive function (Trail Making Test A & B, TMT [46]), anthropometric data (body weight, height, body mass index, waist and hip circumference), body composition (bioelectrical impedance analysis parameters including fat mass, fat-free mass, reactance, resistance, and phase angle), cardiovascular markers (resting heart rate, systolic and diastolic blood pressure, and the ankle-brachial index), laboratory markers (including HbA1c, lipid profile, C-reactive protein, gamma-GT, glucose, hemoglobin, and electrolytes), and physiological outcomes (peak oxygen uptake VO peak, ventilatory thresholds VT1 and VT2, peak power output, maximum heart rate, blood lactate, and blood gases) will be assessed at baseline and after 3 months of intervention (t1).

In addition, various PROMs are collected at baseline, at time point t1 and 12 months post-randomisation (t2). These include physical activity (European Health Interview Survey – Physical Activity Questionnaire, EHIS-PAQ [47]), sleep quality (Pittsburgh Sleep Quality Index, PSQI [48]), health-related quality of life (EuroQol 5-Dimension 5-Level, EQ-5D-5L [49]), depressive symptoms (Patient Health Questionnaire-9, PHQ-9 [50]), loneliness and social isolation (UCLA Loneliness Scale, UCLA-LS [51]), and diabetes risk (Diabetes Risk Test, DRT [52]). Additionally, for the IHHE and IHHE-S groups, blinding success will be evaluated as a separate outcome at t1 [43]. The respective score for each PROM will be calculated and used for analyses. An additional MoCA assessment will also be conducted at t2 to evaluate longer-term effects as secondary outcome. Table 3 provides an overview of all the outcomes alongside their respective survey timepoints.

**Table 3:**
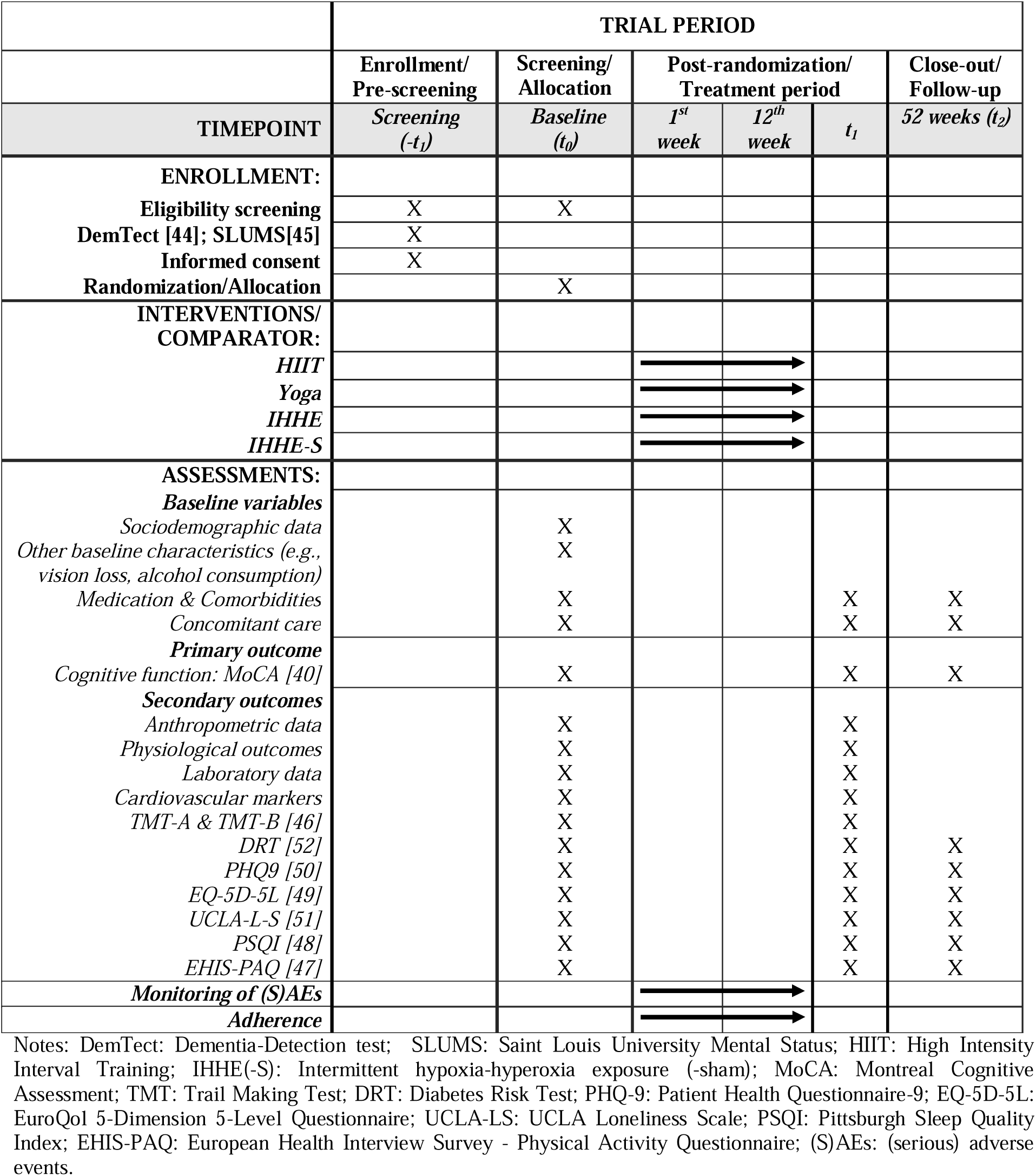
Participant timeline: Schedule of enrollment, interventions, and assessments [29].

|  | TRIAL PERIOD |  |  |  |  |  |
| --- | --- | --- | --- | --- | --- | --- |
|  | Enrollment/<br>Pre-screening | Screening/<br>Allocation | Post-randomization/<br>Treatment period |  |  | Close-out/<br>Follow-up |
| TIMEPOINT | Screening<br>(-t<sub>1</sub>) | Baseline<br>(t<sub>0</sub>) | 1<sup>st</sup><br>week | 12<sup>th</sup><br>week | t<sub>1</sub> | 52 weeks (t<sub>2</sub>) |
| **ENROLLMENT:** |  |  |  |  |  |  |
| Eligibility screening | X | X |  |  |  |  |
| DemTect [44]; SLUMS[45] | X |  |  |  |  |  |
| Informed consent | X |  |  |  |  |  |
| Randomization/Allocation |  | X |  |  |  |  |
| **INTERVENTIONS/**<br>**COMPARATOR:** |  |  |  |  |  |  |
| HIIT |  |  | → |  |  |  |
| Yoga |  |  | → |  |  |  |
| IHHE |  |  | → |  |  |  |
| IHHE-S |  |  | → |  |  |  |
| **ASSESSMENTS:** |  |  |  |  |  |  |
| **Baseline variables** |  |  |  |  |  |  |
| Sociodemographic data |  | X |  |  |  |  |
| Other baseline characteristics (e.g.,<br>vision loss, alcohol consumption) |  | X |  |  |  |  |
| Medication & Comorbidities |  | X |  |  | X | X |
| Concomitant care |  | X |  |  | X | X |
| **Primary outcome** |  |  |  |  |  |  |
| Cognitive function: MoCA [40] |  | X |  |  | X | X |
| **Secondary outcomes** |  |  |  |  |  |  |
| Anthropometric data |  | X |  |  | X |  |
| Physiological outcomes |  | X |  |  | X |  |
| Laboratory data |  | X |  |  | X |  |
| Cardiovascular markers |  | X |  |  | X |  |
| TMT-A & TMT-B [46] |  | X |  |  | X |  |
| DRT [52] |  | X |  |  | X | X |
| PHQ9 [50] |  | X |  |  | X | X |
| EQ-5D-5L [49] |  | X |  |  | X | X |
| UCLA-L-S [51] |  | X |  |  | X | X |
| PSQI [48] |  | X |  |  | X | X |
| EHIS-PAQ [47] |  | X |  |  | X | X |
| **Monitoring of (S)AEs** |  |  | → |  |  |  |
| **Adherence** |  |  | → |  |  |  |

Notes: DemTect: Dementia-Detection test; SLUMS: Saint Louis University Mental Status; HIIT: High Intensity Interval Training; IHHE(-S): Intermittent hypoxia-hyperoxia exposure (-sham); MoCA: Montreal Cognitive Assessment; TMT: Trail Making Test; DRT: Diabetes Risk Test; PHQ-9: Patient Health Questionnaire-9; EQ-5D-5L: EuroQol 5-Dimension 5-Level Questionnaire; UCLA-LS: UCLA Loneliness Scale; PSQI: Pittsburgh Sleep Quality Index; EHIS-PAQ: European Health Interview Survey - Physical Activity Questionnaire; (S)AEs: (serious) adverse events.

### Ancillary studies

As part of an additional study, some subjects will be interviewed in a qualitative study. These will take place after the respective intervention period has ended. A separate information and consent form will be provided. An additional consent form is required for the optional collection of blood samples for further analysis if additional financial resources can be acquired. Furthermore, measurements via functional near-infrared spectroscopy are planned as part of a subproject of the KAYH study. The subjects selected for this subproject will be informed separately, and a separate consent form will be provided. As all three additional assessments are only being conducted as part of individual ancillary studies for which additional funding is required, they will not be discussed further below.

### Safety

Safety monitoring will include continuous supervision during intervention sessions by qualified staff. All harms and (S)AEs will be recorded throughout the intervention phase and assessed systematically at each study visit. Participants will be instructed to report any symptoms or adverse experiences throughout the intervention period, whether or not they consider them related to the intervention. In addition, study staff will actively inquire about adverse events during scheduled assessments and intervention sessions. (S)AEs will be categorized by the project leaders of the three participating institutions together with the principal investigator responsible for the sports medicine baseline diagnostics as intervention-related (confirmed, possible, none). The events will be analyzed descriptively. In the event of an (S)AE, the aforementioned individuals will confer during the intervention regarding the further management of the affected participant and the continuation of the study. Medical clearance will be obtained prior to participation, and predefined stopping criteria will be applied during exercise testing and training to minimize risks. Participants who experience study-related harm will be provided with appropriate medical care and compensation in accordance with institutional policies and applicable regulations. No additional ancillary or post-trial care beyond standard clinical management is planned.

The anticipated harm related to IHHE is expected to be minimal. However, some participants may experience mild and transient adverse effects. These may include discomfort associated with mask wearing, mild dizziness, and sleep disturbances. These adverse events are generally considered non-serious and self-limiting on the basis of literature on IHHE interventions among patients with MCI [36, 37]. Expected harms from the yoga and HIIT interventions may include musculoskeletal discomfort (e.g. soreness or joint strain), mild fatigue, and shortness of breath during exertion. These effects are usually short-lived and resolve without intervention.

Participants will be reminded of scheduled visits for assessments via notes. Flexible scheduling for assessment visits and intervention sessions will be offered to accommodate participants’ availability. To increase participation in follow-up at t2, participants are offered the opportunity to take the assessments digitally. For participants who discontinue or deviate from the intervention protocol, the intention is to systematically record the reasons for withdrawal and collect all available outcome data, provided that the patients provide the necessary information and consent. Apart from the reasons for discontinuing participation, no specific outcomes are recorded for participants who drop out.

## Data collection and management

All outcome assessments will be conducted by trained study staff following standardized operating procedures (SOPs) to ensure accuracy, reliability and consistency. The procedures for all assessments are tested in advance as part of a pilot study and then defined in SOPs. Data collection forms will be made accessible alongside study data via open data sharing (Zenodo: https://zenodo.org).

The MoCA and the DemTect will be administered by trained psychologists or physicians. The DemTect is a brief cognitive screening tool specifically developed to identify MCI and early stages of dementia [44]. Scores range from 0 to 18, with higher scores indicating better performance and thus less cognitive impairment. Scores are adjusted for age and interpreted to indicate normal/age-appropriate cognitive functioning, MCI, or suspected dementia [44, 53]. The primary outcome measurement MoCA is a comprehensive screening tool designed to detect MCI across various domains. The score ranges from 0 to 30 points, with higher scores indicating better cognitive performance. To correct for educational level, individuals with ≤12 years of formal education are awarded one additional point [40]. During the follow-up assessments, it may not always be possible to carry out the MoCA assessment in person at the study centre. In such cases, the assessment can be carried out digitally. The MoCA has demonstrated greater sensitivity than the Mini-Mental State Examination in detecting MCI and early dementia syndromes, and is available and validated in Germany [54]. Additionally, it has already been demonstrated that the MoCA is a reliable and accurate tool for administration via telehealth and that it is well received by participants [55].

The PROMs will be assessed digitally directly via the REDCap platform on a tablet or computer. Standardized, established questionnaires will be used for this purpose. The EHIS-PAQ is used to assess physical activity levels and calculate the HEPA index and the fulfilment of strength recommendations. The questionnaire has been shown to be reliable and valid [56]. The DRT is a standardized self-assessment tool designed to estimate an individual’s risk of developing type 2 diabetes mellitus within ten years and has already been validated in Germany [52, 57]. The PHQ-9 is a self-administered questionnaire used to screen for and assess the severity of depressive symptoms [50]. The construct validity of the PHQ-9 for detecting depression and subthreshold depressive disorder has been shown [58]. The EQ-5D-5L is a standardized measure of health-related quality of life with 5 subscales and a visual analog scale and appears to be a valid measurement instrument [49, 59, 60]. The UCLA Loneliness Scale is a validated and reliable self-report instrument designed to assess subjective feelings of loneliness and social isolation [51, 61]. The PSQI is a self-rated questionnaire that assesses sleep quality and disturbances over a one-month period [48]. Backhaus et al. (2002) reported that the scale has good reliability and validity [62].

Further assessments will be carried out by trained staff in accordance with predefined SOPs. The TMT is a widely used neuropsychological instrument to evaluate visual attention, processing speed, and executive function, and a German normative study has been performed [46]. Anthropometric measurements will be obtained following standardized protocols. Body weight and height will be measured using calibrated scales and stadiometers. Waist and hip circumferences will be measured with non-stretchable tape measures following standardized protocols. For the assessment of body composition, BIA 101 BIVA (Akern Srl, Pontassieve, Italy) will be used. The resting heart rate, blood pressure (systolic and diastolic) and the ankle-brachial index (ABI) will be measured while the participants are in a laying position after a standardized rest period. Venous blood samples will be collected after an overnight fast by trained personnel. This may require an additional visit to the study center in the morning. Cardiorespiratory fitness and exercise-related physiological parameters will be assessed using a ramp test on a cycle ergometer. The test will start at 30 watts, with an increase of 15 watts each minute. Peak oxygen uptake (VO peak) will be determined via spiroergometry and ventilatory thresholds (VT1 and VT2) will be identified [63]. Peak power output (PPO) will be recorded as the highest workload achieved during the test and maximum heart rate (HR_max_) will be measured as the highest heart rate obtained during the test. Blood lactate concentrations and blood gases (e.g., pH, pO_2_, and pCO_2_) will be measured from capillary blood samples collected pre- and post-exercise to assess the metabolic response and exercise intensity domains.

The data collection tool REDCap will be used as the data management platform. REDCap is a free, secure, browser-based software application for designing and managing databases, developed at Vanderbilt University and maintained and distributed under the REDCap Consortium [64, 65]. REDCap can be accessed on mobile devices (smartphones, tablets) via its dedicated application and supports numerous export formats. REDCap has minimal technical requirements and dependencies while providing maximum data security through storage on the secured servers of the host university. The hosting and operation of the web application are self-managed and independent of the REDCap Consortium. Accordingly, the setup and maintenance of REDCap are carried out at the University Hospital Tübingen. Only trained staff will perform data entry. Data range checks will be performed in REDCap using reference values to ensure that data is entered correctly. To minimize the risk of errors during data entry, a random, selective data check will be performed by a researcher not involved in data entry, who will compare the REDCap data with the original data for corresponding outcomes. The items for the PROMs are created as mandatory fields in REDCap to ensure completeness. For paper-based outcomes, SOPs are followed to minimize errors, and data entry in REDCap is also based on mandatory fields, with ranges and plausible values specified to prevent transcription errors.

### Confidentiality

Personal data of the participants will be treated confidentially and will only be accessible to individuals directly involved in the study. The collected personal data and additional medical information will initially be recorded either on paper or in digital form in a pseudonymized manner. Data documented in paper form will subsequently be entered into the secure electronic REDCap study database. A link to personal identity can be made via the participant reidentification list. This list, along with the signed informed consent forms and data protection declarations, will be stored in a locked filing cabinet. Participants may request information on their stored data at any time and have the right to request corrections of inaccurate data. They may also request deletion or anonymization of their data at any time, such that a link to their identity can no longer be established. The collection, storage, use, and transfer of data require explicit consent through the signed data protection consent form. Should participants withdraw from the study, they may decide whether the data already collected should be destroyed or may continue to be used, unless the data has already been included in an analysis or has been published.

Study data will be compiled for joint analysis on the REDCap data platform. To ensure data availability, all study data will additionally be exported into a common, widely used data format. Access to the data will be restricted to authorized study personnel via password-protected computers. All laboratory samples will be stored and analyzed in a pseudonymized form. Only authorized staff will have access to the study data, and all are bound by confidentiality obligations. The data collected in this study may also be used and further processed for future research projects of the departments or institutes in a pseudonymized form. If the participants consent, research data generated within this study will be made publicly available exclusively in anonymized form, free to access, reuse, modify, and distribute without restrictions (open data sharing via Zenodo: https://zenodo.org). The research results from this study will be published in anonymized form in peer-reviewed journals or scientific databases.

## Statistical methods

The baseline characteristics of the study population will be described using descriptive statistics. Continuous variables will be summarized as mean (standard deviation, SD) or median (interquartile range) as well as minimum and maximum (range). Categorical variables will be presented as absolute and relative frequencies. Prognostic factors for clinical endpoints at t1 will be evaluated exploratorily using logistic regression (nominal endpoints) or linear regression (continuous data). Analyses of implementation and patient safety will be descriptive. Data will be analyzed using Excel, IBM SPSS Statistics, and R software.

The primary objective of the analysis is to assess the effectiveness and safety of the three nonpharmacological interventions compared with the sham intervention. The primary outcome measure is the change in the total MoCA score between baseline and post-intervention, compared across intervention groups. The primary analysis of the outcome measures will be performed using a baseline-adjusted analysis of covariance (ANCOVA) as a function of group allocation at t1 (α = 0.05, two-sided). For the primary endpoint, a closed testing procedure will be applied to conduct pairwise comparisons between the four treatment groups, control the overall type I error rate, and maintain the global significance level at α = 5% (two-sided). Initially, a global test will be performed across all four treatment groups to test the null hypothesis of equal means. If this null hypothesis is rejected, three pairwise comparisons (HIIT vs. sham, yoga vs. sham, and IHHE vs. sham) will be tested confirmatorily. Comparisons among the three active interventions will be tested exploratorily. In addition to p-values, 95% confidence intervals will be reported. To assess the clinical relevance of the results, effect sizes for between-group comparisons will be calculated. The analysis of secondary outcomes and time points will follow the same approach as for the primary analysis. However, only the primary endpoint will be analyzed confirmatorily, as no adjustment for multiple testing is planned. The primary analysis population will be the intention-to-treat (ITT) population, defined as all individuals with baseline values for the primary endpoint who have attended at least one intervention session. Sensitivity analyses will include a complete case analysis (analysis of participants with available data) and a per-protocol analysis, including only those who have completed at least 75% of the planned intervention sessions. For the primary analysis population (ITT), missing data will be imputed using multiple imputation if more than 5% of the data are missing and/or if a systematic selection bias among drop-outs is suspected.

The same statistical methods applied in the primary analysis will be used for sensitivity analysis. The planned ancillary studies (further analyses of blood samples, functional near-infrared spectroscopy, and qualitative interviews) will only examine subgroups. Participants who are interested will be examined and analyzed accordingly. If the acquisition of appropriate funding is successful, detailed analyses for the subgroups of each sub-study will be determined within the framework of the respective studies. To evaluate the data from time point t1 within the funding period, an interim analysis is performed on time point t1 for all participants. The analysis will be carried out as soon as all participants have completed time point t1 and the relevant data is available.

## Oversight and monitoring

The trial is coordinated by a coordination center led by one of the three involved institutions, which assumes overall responsibility for the day-to-day management and the operational oversight. This team provides continuous organizational support throughout the study, with regular meetings held on a monthly basis to monitor progress, address operational challenges, and ensure adherence to the protocol. A Trial Steering Committee has not been established for this study. Oversight and strategic decision-making responsibilities are managed by the principal investigators of the three involved institutions. Additional committees, such as Data Monitoring Committee, are not convened. Participant safety and endpoint assessment are managed within the study team. No Data Monitoring Committee will be established. Safety monitoring and oversight will be conducted by the principal investigators and the study team according to standard procedures. The central study monitoring will be performed by members of the study team who are independent of the enrollment and data entry process to ensure objectivity. Monitoring activities are scheduled approximately every three weeks as part of regular audits. No audit trial will be conducted. However, central study monitoring includes a thorough check of consent forms and adherence to eligibility criteria, as well as random, selective checks on the plausibility of data entry.

## Ethics and dissemination

This study has been approved by the Ethics Committee of the Medical Faculty of the University of Tübingen (494/2025BO1). Written, informed consent to participate will be obtained from all participants. In the case of modifications of the protocol, an amendment will be submitted for review and approval by the responsible local ethics committee. Any modifications will also be updated in the trial registry to ensure transparency, and the external funder will be informed. The findings of this study will be disseminated through publication in peer-reviewed journals and presentation at scientific conferences and to relevant stakeholders.

## Discussion

Dementia is a growing global health concern, with increasing prevalence and substantial impacts on healthcare systems and society as a whole. Early interventions in individuals with MCI represent a promising strategy for dementia prevention. Currently, no curative pharmacological therapies for MCI exist, and nonpharmacological approaches remain insufficiently investigated. Therefore, this randomized sham-controlled trial aims to evaluate the effects of HIIT, yoga, and IHHE on cognitive function among people with MCI.

A shared underlying mechanism across the three active intervention arms is the sustained improvement of cerebral blood flow and mitochondrial function through changes in cerebral oxygen availability [13, 21, 26]. In this context, this study could provide further evidence of the mechanisms and effects involved. Furthermore, this study has the potential to provide valuable, practice-relevant information on the efficacy, safety, and feasibility of these nonpharmacological interventions for conditions with currently limited therapeutic options and no approved pharmacological treatments. We hypothesize that, compared with a sham control, HIIT, yoga, and IHHE will demonstrate superiority in the enhancement of cognitive function. The findings of this study may provide a basis for future therapeutic interventions targeting individuals with MCI, with the aim of delaying or preventing the onset of dementia. Overall, this study is expected to provide valuable insights that could guide clinical decision-making and contribute to the broader scientific knowledge in this field. The results will be disseminated through peer-reviewed publications and presentations in accordance with open and transparent research practices. The planned sub-studies (further analyses of blood samples, functional near-infrared spectroscopy, and qualitative interviews) could also provide further insights into the mechanisms of the interventions. In addition to analyzing specific responders, it will also be possible to capture the subjective experiences of the participants. These sub-studies may serve as a foundation for subsequent research.

## Strengths and limitations

A major strength of this study is the randomized, sham-controlled design, which allows for robust comparisons between intervention groups and control group. The inclusion of a sham group helps to control for potential placebo effects. Furthermore, the planned sample size is adequately powered to detect meaningful differences in cognitive outcomes among patients with MCI. The follow-up assessment 12 months after the baseline visit allows the examination of the long-term effects of the interventions. However, there are several limitations to consider. Owing to the nature of the interventions, blinding of participants and therapists is not feasible for all intervention groups. Additionally, PROMs may be subject to reporting bias or variability in responses from participants.

## Trial status

Recruitment will begin in January 2026 and will be completed by approximately December 2026.

## Data Availability

All data produced in the present work are contained in the manuscript

## Abbreviations

ABI: Ankle-brachial index
BIA: Bioelectrical impedance analysis
DemTect: Dementia detection test
DRT: Diabetes risk test
EHIS-PAQ: European Health Interview Survey – Physical Activity Questionnaire
EQ-5D-5L: EuroQol 5-Dimension 5-Level Questionnaire
FiO_2_: Fraction of inspired oxygen
FITT: Frequency, intensity, time and type
HIIT: High intensity interval training
IHE: Intermittent hypoxia exposure
IHHE: Intermittent hypoxia-hyperoxia exposure
IHHE-S: Intermittent hypoxia-hyperoxia exposure sham
IHHT: Intermittent hypoxia-hyperoxia training
KAYH: Aerobic interval training, yoga, hypoxia exposure to improve cerebral oxygenation in mild cognitive impairment
MCI: Mild cognitive impairment
MoCA: Montreal Cognitive Assessment
PHQ-9: Patient Health Questionnaire-9
PROMs: Patient-reported outcome measures
PSQI: Pittsburgh Sleep Quality Index
REDCap: Research Electronic Data Capture
SLUMS: Saint Louis University Mental Status
SOP: Standardized operating procedure
SPIRIT: Standard Protocol Items: Recommendations for Interventional Trials
SpO: Peripheral oxygen saturation
TIDieR: Template for Intervention Description and Replication
TMT: Trail making test
UCLA-LS: UCLA Loneliness Scale
VO_2_ peak: Peak oxygen uptake

## Declarations Acknowledgements

We acknowledge support by Open Access Publishing Fund of University of Tübingen.

## Authors’ contributions

HC, GE and IK are the principal investigators and conceived the study, and led the proposal and protocol development. All authors contributed to study design and to development of the proposal. SW provided the first draft of the manuscript. All authors provided feedback and suggestions for this manuscript and read and approved its final version.

## Funding

This work was supported by the Karl and Veronica Carstens Foundation (Am Deimelsberg 36, 45276 Essen, Germany) grant number KVC 0/162/2025. The funding body has no role in the study design, the collection, analysis, or interpretation of the data, or in writing the manuscript.

## Competing interests

The authors declare that they have no competing interests.

## Patient and public involvement

Patients and/or the public were not involved in the design, conduct, reporting, or dissemination plans of this research. Research findings will be presented to stakeholders and published in peer- reviewed journals and at conferences.

## Consent for publication

Not applicable.

## Availability of data and materials

If the participants consent, the research data generated within the framework of this study will be made publicly available exclusively in anonymized form, free to access, reuse, modify, and distribute without any restrictions (open data sharing via Zenodo: https://zenodo.org).

## Notes

### Competing Interest Statement

The authors have declared no competing interest.

### Clinical Trial

DRKS00038438

### Author Declarations

Ethics committee of the Medical Faculty of the University of Tuebingen gave ethical approval for this work

